# School-located influenza vaccination and community-wide indirect effects: reconciling mathematical models to epidemiologic models

**DOI:** 10.1101/2022.10.08.22280870

**Authors:** Nimalan Arinaminpathy, Carrie Reed, Matthew Biggerstaff, Anna Nguyen, Tejas S. Athni, Benjamin F. Arnold, Alan Hubbard, John M. Colford, Art Reingold, Jade Benjamin-Chung

## Abstract

**Background:** Mathematical models and empirical epidemiologic studies (e.g., randomized and observational studies) are complementary tools but may produce conflicting results for a given research question. We used sensitivity analyses and bias analyses to explore such discrepancies in a study of the indirect effects of influenza vaccination.

**Methods:** We fit an age-structured, deterministic, compartmental model to estimate indirect effects of a school-based influenza vaccination program in California that was evaluated in a previous matched cohort study. To understand discrepancies in their results, we used 1) a model with constrained parameters such that projections matched the cohort study; and 2) probabilistic bias analyses to identify potential biases (e.g., outcome misclassification due to incomplete influenza testing) that, if corrected, would align the empirical results with the mathematical model.

**Results:** The indirect effect estimate (% reduction in influenza hospitalization among older adults in intervention vs. control) was 22.3% (95% CI 7.6% – 37.1%) in the cohort study but only 1.6% (95% Bayesian credible intervals 0.4 – 4.4%) in the mathematical model. When constrained, mathematical models aligned with the cohort study when there was substantially lower pre-existing immunity among school-age children and older adults. Conversely, empirical estimates corrected for potential bias aligned with mathematical model estimates only if influenza testing rates were 15-23% lower in the intervention vs. comparison site.

**Conclusions:** Sensitivity and bias analysis can shed light on why results of mathematical models and empirical epidemiologic studies differ for the same research question, and in turn, can improve study and model design.

## Introduction

Mathematical modelling offers a helpful tool for capturing the complexities of infectious disease transmission. As well as being applied widely as part of the COVID-19 response,^1–3^ modelling continues to play an important role in planning the response to other major infections.^4^ However, no mathematical model can be fully predictive: all models must make simplifications of reality in order to be tractable, and models may produce different results, even when applied to the same question.^5,6^ Although well-documented,^7,8^ it remains important for these and other limitations to be fully appreciated in any application of mathematical modelling for public health decision-making.

Given such limitations, there can be substantial value in comparing mathematical model projections with estimates from epidemiologic field studies that aim to quantify the impact of interventions on transmission using real-world data. Epidemiologic studies may better capture complex real-world transmission dynamics, yet they are subject to different assumptions and biases than mathematical models. Comparisons of results from the two approaches could shed light on the influence of each approach’s assumptions and biases, and in turn, improve model and study design and cast new light on evidence about intervention effectiveness. Here, we present results of one such analysis, in the context of an intervention to improve rates of influenza vaccination amongst schoolchildren in California, USA.

Vaccination is critical for the prevention of seasonal influenza morbidity and mortality, particularly among vulnerable groups such as the elderly.^9^ In the USA and elsewhere, recommendations for seasonal influenza vaccination extends to all age groups^10^ older than six months of age. Mathematical modelling has highlighted the potential importance of vaccinating schoolchildren for reducing community-wide transmission.^11^, and studies from Japan have provided empirical evidence for such impact.^12^ In this context, the ‘Shoo-the-Flu’ intervention aimed to increase influenza vaccination coverage amongst elementary children in Oakland, California.^13^ Notably, this study aimed to quantify indirect (transmission-mediated) effects: comparison with a closely-matched control district, West Contra Costa, showed evidence of indirect effects through a reduction in influenza-related hospitalizations among non-elementary school aged children and the elderly in Oakland.

We fit a mathematical model to capture these indirect effects. We describe discordance between the mathematical model and empirical epidemiological study and report the results of sensitivity and bias analyses to identify possible reasons for this discordance. We performed analyses in two directions: one to identify influential parameters of the mathematical model, and conversely to identify potential biases in the epidemiological analysis. This study demonstrates how to use sensitivity analysis and probabilistic bias analysis to investigate different results produced by epidemiologic models and mathematical models. We conclude with recommendations for future studies.

## Methods

### 1. Overview of study population and intervention

The mathematical model and prior cohort study^13^ aimed to estimate impact of a city-wide school-located influenza vaccination program on community-wide influenza hospitalization incidence. The Shoo the Flu program (https://www.shootheflu.org) was delivered in Oakland, California, offering delivery of free influenza vaccinations at elementary schools (kindergarten through grade 5). The intervention was initiated in 2014; here, we focused on the 2017-18 season, which had the highest incidence of influenza, and the largest estimate of intervention effectiveness in the cohort study. In this season, the program delivered quadrivalent inactivated influenza vaccinations to 7,536 of 34,741 eligible students in 95 elementary schools. The cohort study enrolled 34 schools in Oakland Unified School District and 34 schools in West Contra Costa Unified School District (the comparison group). Intervention and comparison schools were pair-matched by pre-intervention school-level characteristics (mean enrollment, class size, parental education, academic performance index scores, California standardized test scores, school-level percentage of English language learners, and school-level percentage of students receiving free lunch at school) using a multivariate genetic matching algorithm.^14^ The study conducted a survey of parents and guardians of students to estimate influenza vaccination coverage. The study estimated indirect effects using data on laboratory-confirmed influenza hospitalizations among residents of the zip codes that overlapped with the boundaries of each school district from the CDC-sponsored California Emerging Infections Program (CEIP). The study estimated that vaccination coverage in 2017-18 in children aged 5-11 years was 11 percentage points (95% CI 7%, 15%) higher and influenza hospitalization incidence was 22% (95% CI 8% – 37%) lower in adults 65 years or older in the intervention vs. comparison site.^13^ Additional details on the intervention and cohort study design have been reported elsewhere.^13^

### 2. Mathematical modelling

We modelled the indirect effect of the intervention among adults ≥65 years of age using an age-structured, deterministic, compartmental model, illustrated schematically in Figure 1. We focused on influenza A, which was the predominant influenza type during the 2017/18 season in California. For simplicity, we did not distinguish H3 and H1 subtypes of influenza A. The model was stratified into six different age groups: <4yo, 5-11yo, 12-17yo, 18-49yo, 50-64yo, and ≥65yo. To capture mixing between different age groups, we drew from contact matrices recently estimated for the USA.^15^ The overall structure shown in Figure 1 was further stratified by influenza vaccination status, distinguishing those who did and did not receive seasonal influenza vaccination during the 2017/18 season. Given the focus of this analysis on indirect effects, we assumed that the measured vaccine efficacy is against infection and, moreover, that vaccination confers protection through a ‘leaky’ mechanism: that is, we assumed that all receiving influenza vaccination have their risk of infection reduced by an amount equivalent to the vaccine efficacy. Because the majority of seasonal vaccination coverage in the USA is typically completed prior to the onset of the influenza season^16^, we modelled age-specific vaccination simply through initial conditions, ensuring that the initial population vaccinated compartments reflected the estimated, age-specific vaccination coverage.

**Figure 1.**
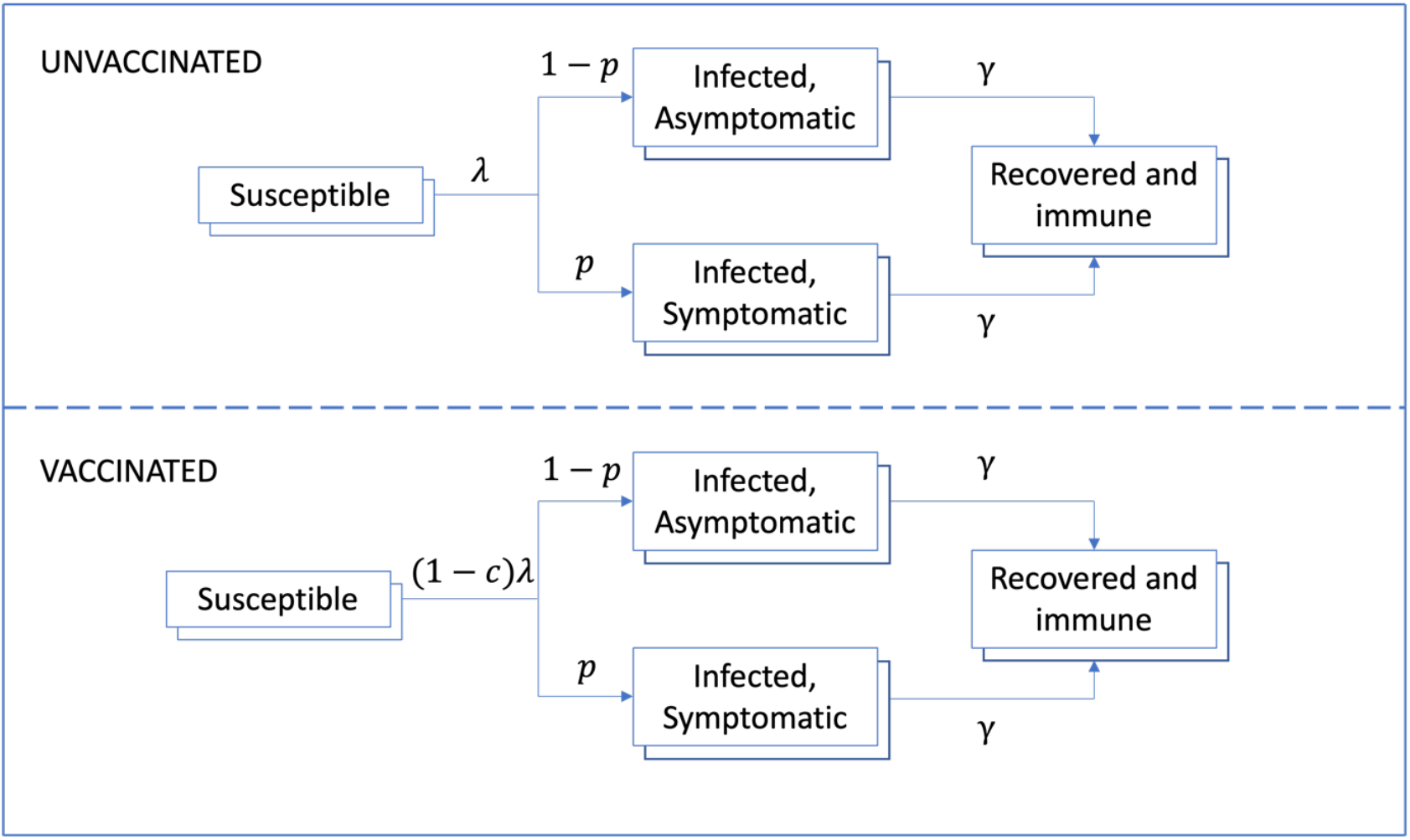
Schematic illustration of the mathematical model. We distinguish those who were vaccinated in time for the 2017/18 season (lower panel) from those who were not (upper panel). Model symbols are as follows: force-of-infection (λ), vaccine efficacy in reducing the force-of-infection (*c*), proportion of infections that are symptomatic (*p*), and per-capita recovery rate (*γ*). ‘Layers’ to each compartment denote stratification into six age groups: <4yo, 5-11yo, 12-17yo, 18-49yo, 50-64yo, and ≥65yo.

Since data on pre-intervention influenza vaccination coverage by site were not available, we drew from state-level estimates of vaccination coverage in California, assuming coverage was the same in the intervention and control districts (see Fig. S1A) (C. Reed, personal communication, 31 Aug 2020). We drew from national-level estimates of vaccine efficacy against influenza A for the 2017/18 season (see Fig. S1B).

To model the intervention, we simulated the impact (reduction in hospitalizations amongst those aged ≥ 65 yo) of increasing vaccination in the 5-11yo age group from 58% to 69%. Although in theory the intervention may have also influenced other age groups to increase uptake of vaccination, we had no data to this effect: thus, we assumed no change in vaccination coverage in other age groups.

#### Calibration data

We calibrated model parameters in order to match the available epidemiological data in the control district (West Contra Costa). In particular, as a proxy for incidence, we used district-level data from the CEIP for age-specific, weekly hospitalizations that were virologically confirmed as being influenza. These are the same data that were used to estimate indirect effects in the cohort study. To link these data with the weekly incidence of symptomatic influenza in the community, we drew from previous estimates by CDC for the age-specific proportions of symptomatic influenza cases that are hospitalized, tested for influenza, and reported to FluSURV-NET.^17^ These multipliers are available by age group, but only at the national level. Further details on model calibration are provided in Supplement 1.

Finally, using a sensitivity analysis, we sought to identify model parameters that would best explain differences in the results of the transmission model and the cohort study. To do so, we performed an alternative, ‘constrained’ calibration, one where – in addition to the data described above – we also included the requirement that model projections should capture the observed reduction in hospitalization in those ≥ 65 years of age.

### 3. Examining potential sources of error in study data

Focusing next on the statistical analysis of the study data, using probabilistic bias analysis,^18^ we sought to identify sources of bias that, if corrected, would result in cohort study estimates that aligned with those of the mathematical model. We explored the following bias scenarios, which we considered plausible for the cohort study: 1) incomplete influenza testing among hospitalized patients, and 2) an unmeasured time-dependent confounder of the relationship between the school-located influenza vaccination intervention and influenza hospitalization.

#### Correction for potential misclassification of influenza hospitalization

Our original study estimated impacts of school-located influenza on laboratory-confirmed influenza hospitalization. If some patients who truly had influenza were not tested for influenza, this could result in outcome misclassification, and if testing rates differed between the intervention and comparison sites, this could result in differential outcome misclassification. We corrected estimates of intervention impact on hospitalization for influenza on potential misclassification due to incomplete influenza testing. We obtained estimates of the proportion of hospitalized patients with pneumonia and influenza International Statistical Classification of Disease codes who were tested for influenza from the CEIP, which provided influenza hospitalization data in the original study. These unpublished data were collected as part of CEIP’s evaluation of its activities in Alameda, San Francisco, and Contra Costa counties. It was not possible to stratify data by study site. First, we performed a probabilistic bias analysis assuming that the proportion of patients ≥ 65 years of age that were tested for influenza was 0.58 on average and ranged from 0.38 to 0.78 (Beta(α=44, β=32)); we defined the prior based on testing rate estimates from CEIP and used the same prior distribution for the intervention and comparison sites (Table S2). Next, to explore the influence of different testing rates in the intervention and comparison sites, we corrected case counts for all combinations of testing values from the prior distribution for each site in increments of 0.01. We corrected the number of hospitalized cases in each study site for incomplete influenza testing using the following formula:

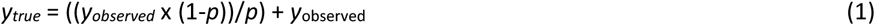

where *y*_*true*_ is the number of cases hospitalized with laboratory-confirmed influenza corrected for incomplete testing, *y*_*observed*_ is the observed number of cases, and *p* is the proportion of hospitalized patients tested for influenza.

#### Correction for potential unmeasured time-dependent confounding

We hypothesized that the implementation of the main provisions of the Affordable Care Act (ACA), which coincided with the start of the intervention in 2014, could have changed health insurance coverage over time differentially between sites. This may have introduced time-dependent unmeasured confounding that would not have been removed by the original difference-in-difference analysis. We define *Y* as the outcome, *X* as an indicator for the intervention vs. comparison site, and *C* as an indicator for the unmeasured confounder (health insurance coverage). Priors for correcting for an unmeasured confounder include: 1) the probability of the confounder in the intervention group P(*C*|*X*=1), 2) the probability of the confounder in the comparison group P(*C*|*X*=0), and 3) difference in risk between hospitalization (*Y*) and the confounder (*C*) in the comparison group (*X*=0), assuming no effect modification by *X* (*E*[*Y*|*C* = 1, *X* = 0] − *E*[*Y*|*C* = 0, *X* = 0]).^18^ We defined priors separately for the period before and during the intervention to allow for time-dependent confounding. We defined both realistic and alternative, less realistic priors to investigate what priors would be required to replicate the mathematical model’s findings (see details in Supplement 2).

We calculated three Mantel-Haenszel risk differences (RD_MH_) correcting for confounding: 1) the risk difference for the intervention in the pre-intervention period (RD_MH, pre_), 2) the risk difference for the intervention during the intervention (RD_MH, post_), and 3) the risk difference comparing the risk during the intervention vs. prior to the intervention in the comparison group (RD_MH, comparison_).^18^ We calculated the difference-in-difference correcting for bias as RD_MH, post_ - RD_MH, pre_ and the relative reduction in the difference-in-difference correcting for bias as (RD_MH, post_ - RD_MH, pre_)/ RD_MH, comparison_.

In all bias analyses, we randomly sampled bias parameters from prior distributions without replacement and repeated analyses 10,000 times to obtain distributions of bias-corrected estimates. We used a bootstrap with 1000 replicates to obtain credible intervals for the ratio of mean prior values for each prior.

## Results

### Mathematical model estimates

Figure 2 shows results of model calibration for the observed epidemic in West Contra Costa, the ‘control’ district. Despite the scarcity of data in the three youngest age groups, the model captures a reasonable fit with the dynamics of influenza hospitalization in the three oldest age groups. Figure S2 shows model fits to cumulative numbers hospitalized, again showing a reasonable fit, while Figure S3 illustrates the marginal posterior distributions for each of the model parameters.

**Figure 2.**
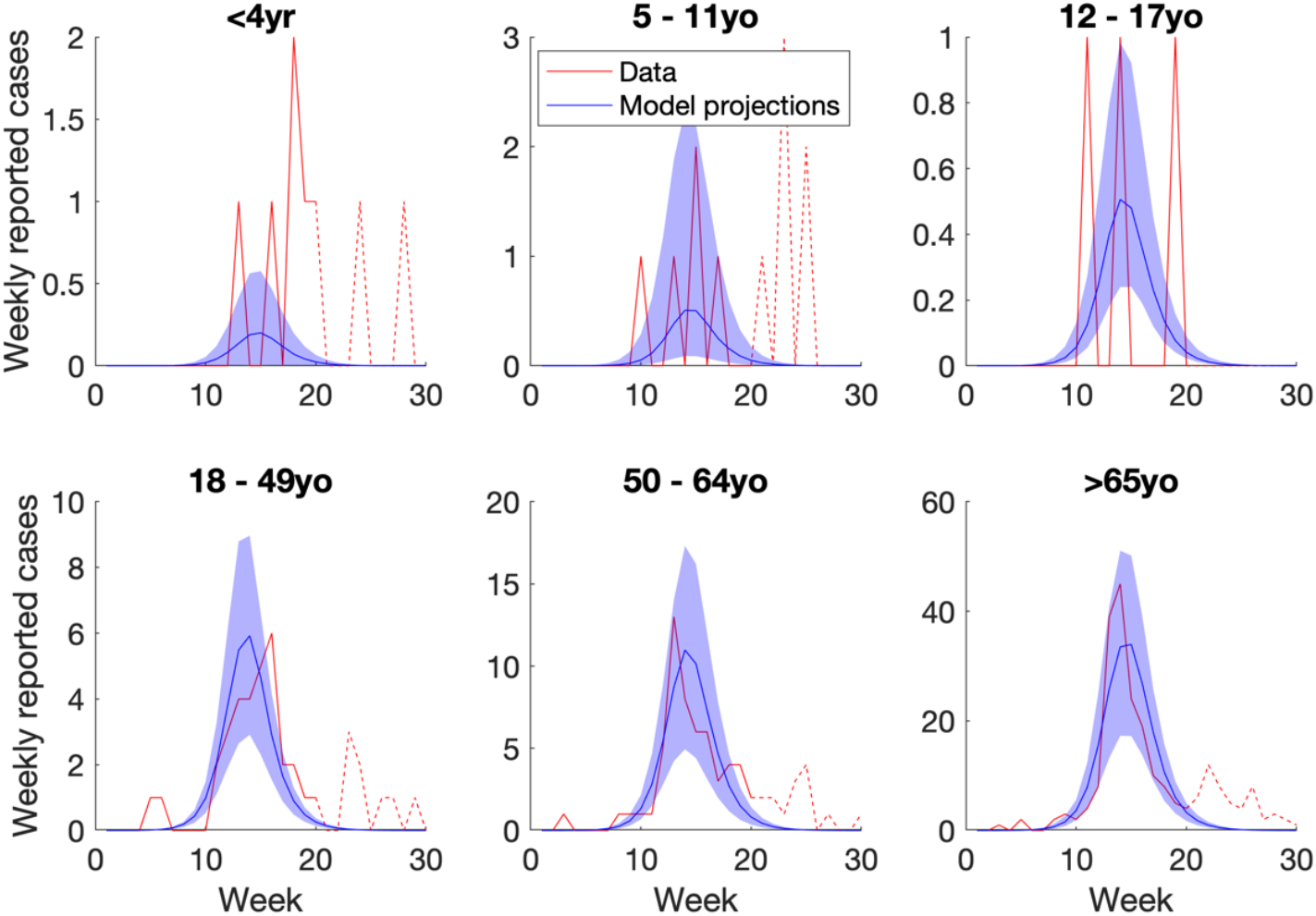
Results of model calibration to epidemiological data in the control district (West Contra Costa). Curves in red show age-specific data on virologically confirmed influenza hospitalizations from FluSURV-NET. Solid red lines show parts of the epidemic when influenza A dominated (to which the data were calibrated) while dashed red lines show parts of the epidemic driven by influenza B (not addressed in the calibration). Blue curves show best model fits, scaled by multipliers associating the data with incidence of symptomatic influenza, with shaded regions showing 95% Bayesian uncertainty intervals. While this Figure shows model fits to the weekly data, Figure S2 in the supporting information also shows model fits to cumulative reported cases.

We next simulated the effects of an intervention to increase influenza vaccination coverage in schoolchildren 5-11 years old, by 11 percentage points (i.e. increasing coverage from 58% to 69%). Table 1 shows the reductions in hospitalizations that were projected to result in each of the other age groups. Notably, model projections suggest a 1.6% reduction (95% Bayesian credible intervals 0.14 – 4.4%) in hospitalizations in those ≥ 65 years of age, substantially lower than the empirically observed reduction of 22%.

**Table 1.** Model-simulated indirect effects. (reduction in hospitalizations in those ≥65 of age) arising from increasing vaccination in schoolchildren

| **Model type** | **Percent reduction in cumulative hospitalizations in those over 65 years old** |
| --- | --- |
| Unconstrained | 1.6% (Bayesian credible intervals 0.14 – 4.4%) |
| Constrained | 21.7% (Bayesian credible intervals 20.0 – 23.3%) |

To examine which parameters are most strongly associated with this low impact, we performed the ‘constrained’ calibration described above. Figures S3 – S5 show the results of this calibration, in terms of parameter estimates and agreement with the epidemic dynamics. Table 1 illustrates that this model indeed approximates the expected indirect effect in those ≥ 65 years of age.

Figure 3 shows results of a comparison of posterior samples in the unconstrained and constrained calibrations, plotting the ratio between the two. Parameters whose uncertainty intervals cross 0 on the logarithmic axis are those for which there appears to be no systematic difference between constrained and unconstrained posterior densities. Amongst the parameters that do show a systematic difference, two notable examples are: the proportion initially immune in those aged 5-11 years, and in those ≥ 65 years of age. In particular, in order to capture the impact of vaccination, the constrained calibration systematically estimates these parameters as having lower values than in the unconstrained calibration: that is, the mathematical model requires lower pre-existing immunity in both of these age groups in order to capture indirect effects in ≥ 65-year-olds. Amongst other parameters showing a systematic difference between the samples are: the rate-of-infection (systematically estimated as higher in the constrained model); the average duration of infectiousness (lower in the constrained model); and the proportion symptomatic (higher in the constrained model).

**Figure 3.**
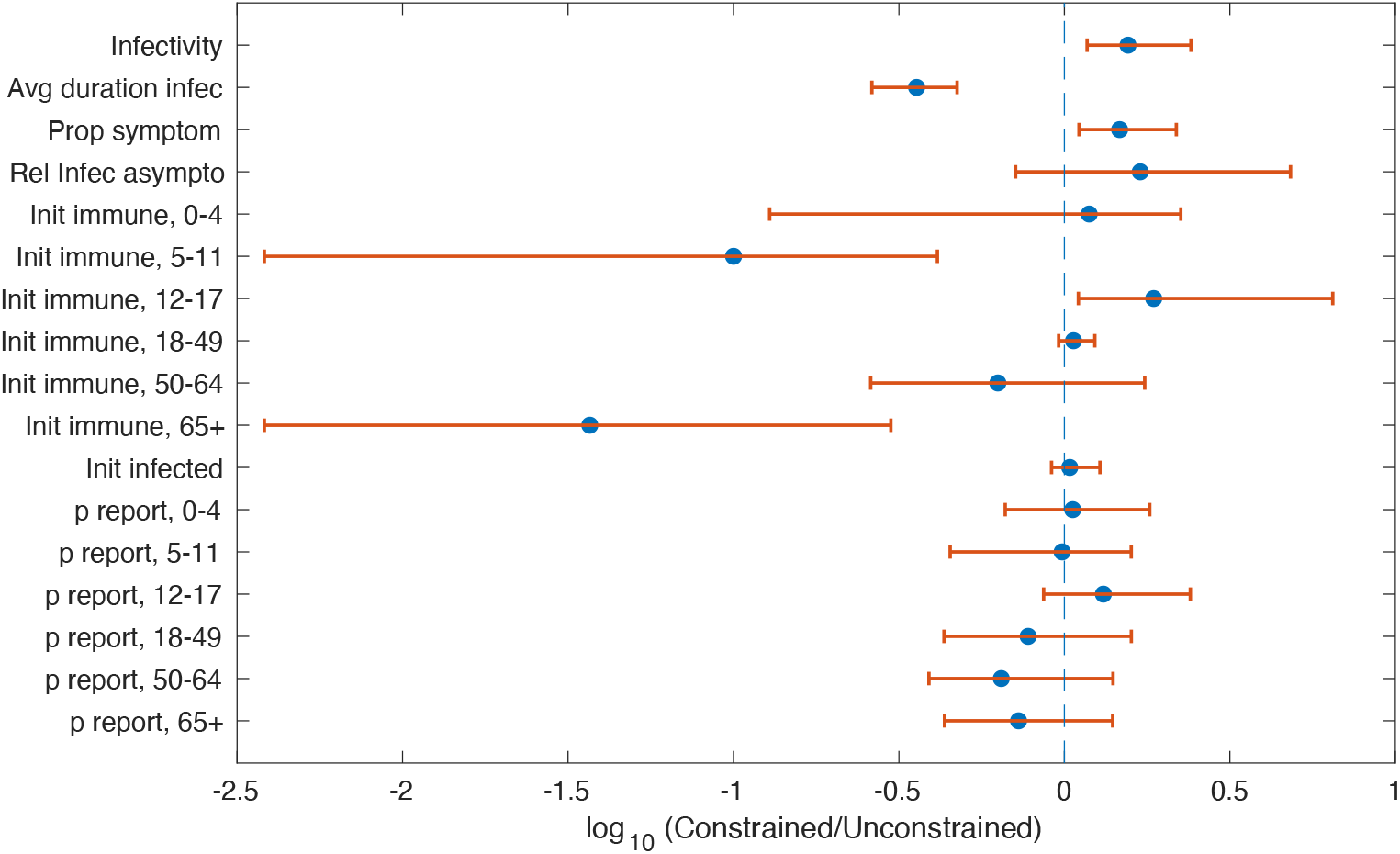
Comparison of parameter estimates between ‘constrained’ and ‘unconstrained’ calibrations. In constrained calibration, we incorporated in the posterior density a likelihood term for the impact of vaccination in those ≥65 years of age. Figure shows estimates for the ratio of parameter values, calculated as constrained vs unconstrained. The vertical dashed line corresponds to a ratio of 1 (hence a log-ratio of zero); uncertainty intervals show 95% Bayesian credible intervals.

### Examining potential sources of error in study data

#### Estimate of intervention impact adjusting for incomplete influenza testing

In the original study, the difference-in-difference comparing influenza hospitalizations in the intervention and comparison areas among adults ≥65 years was −160 per 100,000 (95% CI −267, −53) in 2017-18. Using probabilistic bias analysis to correct for misclassification due to incomplete influenza testing of patients hospitalized for pneumonia and influenza, the mean of difference-in-differences across replications was −276 per 100,000 (range: −412 to −209 per 100,000). No results aligned with the mathematical model. In our exploratory analysis investigating all possible combinations of priors in each site, 1.4% of results aligned with the mathematical model, which estimated a 1.6% reduction in influenza hospitalization in the intervention vs. comparison area, accounting for pre-intervention differences between sites. Bias-corrected relative reductions were similar to the mathematical model (2% to 0%) when influenza testing was approximately 15-23% lower in the intervention site than in the comparison site and when the percentage tested for influenza in the intervention site ranged from 38% to 56% (Figure 4a). The intervention was associated with increased influenza hospitalizations when the percentage tested for influenza was more than 23% lower in the intervention than in the comparison site (Figure 4b). Conversely, the intervention was associated with a larger reduction in influenza hospitalizations than our original analysis when the percentage tested for influenza was higher in the intervention than the comparison site.

**Figure 4.**
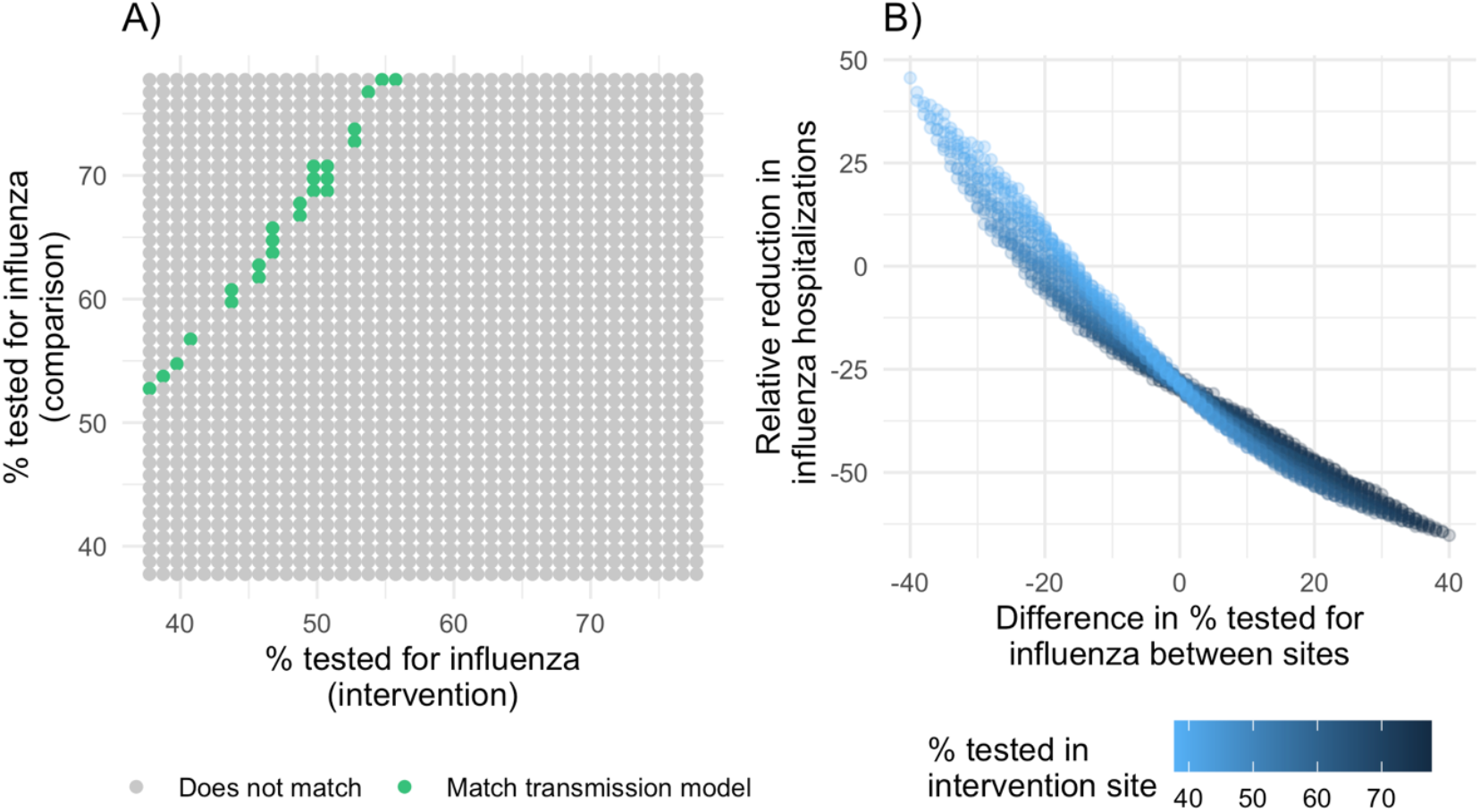
Concordance of epidemiologic and mathematical models by % of patients tested for influenza in the intervention vs. comparison site. Using probabilistic bias analysis, we corrected empirical estimates of the incidence of laboratory-confirmed influenza hospitalizations in the study site for misclassification due to potential incomplete influenza testing of patients hospitalized for pneumonia and influenza. In an exploratory analysis, we investigated all possible combinations of priors in each site. A) shows whether bias-corrected estimates matched empirical estimates by the percentage of patients tested for influenza in the intervention and comparison site. B) shows relative reductions in influenza hospitalization incidence by the percentage of patients tested for influenza in the intervention site and the difference in the percentage tested between sites.

#### Estimate of intervention impact adjusting for unmeasured confounding

After adjusting for potential unmeasured time-dependent confounding with realistic priors, results were similar to the original analysis; the mean of difference-in-differences across replications was −160 per 100,000 (range: −183 to −138 per 100,000), and the mean percentage change was −29% (range: −33% to −25%) (Table 2). No estimates aligned with the results of the mathematical model.

**Table 2.** Percentage change in influenza hospitalizations corrected for time-dependent confounding.

|  | N simulations with valid case counts* | Mean | Range |
| --- | --- | --- | --- |
| Realistic scenario | 10,000 | -29% | -33, -25% |
| Extreme values of health insurance coverage by site | 10,000 | -27% | -114, 78% |
| Extreme values of the relationship between hospitalization and health insurance coverage | 3,615 | -28% | -37%, -20% |

\* Excludes simulations that resulted in zero case counts

In the alternative analysis with extreme prior values of health insurance coverage by site, no results aligned with the mathematical model, and the mean percentage change was −27% (range: −114% to 78%). One percent of bias-corrected estimates aligned with the mathematical model; among those that did, the prior for the difference in risk between hospitalization and the confounder in the comparison group was always closer to zero in the pre-intervention period (RD range: −0.0007, 0.0007) compared to during the intervention period (RD range: −0.0025, −0.0010) (Figures S6-7). This implies that the epidemiologic analysis matched the mathematical model results only under extreme shifts in insurance coverage during the intervention period (from 0 to 90% in the intervention site and from 90% to 0% in the comparison site) and when the effect of health insurance coverage on influenza hospitalizations was weaker prior to compared to during the intervention period. In the alternative analysis with stronger relationships between hospitalization and health insurance coverage, no results aligned with the mathematical model, and the mean percentage change was −28% (range: −37% to −20%).

## Discussion

This study demonstrates how modelling sensitivity analysis and probabilistic bias analysis can be used to examine differences in results from methodologically distinct analyses. While the original study^13^ found that school-located influenza vaccination was associated with a 22.3% relative reduction in community-wide influenza hospitalizations among adults ≥65 years old, a mathematical model that posed the same question using the same underlying data found a relative reduction of only 1.6%. Sensitivity analyses of mathematical models showed that model parameters that would replicate the original study results would require unrealistically low levels of pre-existing immunity to influenza among school-age children and older adults. Other factors included a higher rate of influenza infection, a lower average duration of influenza infection, and a higher proportion of symptomatic individuals. Using probabilistic bias analysis, we identified differential influenza testing rates among hospitalized patients as the most plausible potential source of bias that, if truly present, could explain the discrepancy between the epidemiologic and mathematical models. Considering the relative strengths and weaknesses of each approach, the intervention likely produced indirect effects among older adults, but this study suggests that true effects may have been smaller than estimates published in the original study.

On the modelling side, our analysis yielded markedly more modest impact than previous modelling studies of vaccinating schoolchildren against influenza. These differences are likely due to the comparators involved: a 2009 study in the USA^19^ and a 2013 UK study^11^ both modelled an increase in vaccination coverage amongst children from 0% to 70%. Vaccination coverage in both the USA and UK has increased substantially in recent years: in the present study, we modelled an increase in influenza vaccination coverage in schoolchildren from 58% to 69%. High levels of pre-existing immunity in schoolchildren would tend to reduce the incremental impact of additional vaccination coverage in this group. Likewise, high levels of pre-existing immunity in the elderly would reduce the extent to which they can benefit from indirect effects of vaccination in other age groups. Contrary to the low levels of immunity required by the constrained model, studies in the USA and elsewhere have shown the presence of antibodies to influenza in over 40% of school-aged children.^20–22^ Although some of these studies were performed in the context of the 2009 influenza pandemic, we might expect the prevalence of antibodies to seasonal influenza to be comparable or greater, arising from several seasons of exposure to seasonal influenza viruses as well as vaccination in past seasons.

Epidemiologic analyses aligned with the mathematical model only when the percentage of patients ≥ 65 years old who were tested for influenza was 15-23% lower at hospitals in the intervention area vs. comparison area. While it is not possible to verify testing rates by site, we consider this difference in testing rates to be plausible because it is within the range of observed testing rates, which varied from 21-92% depending on the provider type and age group during the study period. The California Department of Public Health mandates reporting of laboratory-confirmed influenza deaths among individuals 0-64 years of age, but not influenza hospitalizations. Whether a physician orders an influenza test for a patient likely depends on a range of factors that could plausibly vary between study areas, including hospital protocols, laboratory capacity, patient insurance type, patient risk factors, patient symptoms, and the severity of circulating influenza strains at a given time.

The analysis for possible unmeasured time-dependent confounding due to the start of the ACA did not explain the discrepancy in the mathematical model and the epidemiologic study’s findings. Confounding-corrected estimates aligned with mathematical models only when we assumed extreme, unrealistic changes in insurance coverage that were in the opposite direction between study sites. Most studies have not found evidence that the ACA led to increased influenza vaccination,^23–26^ and CDC estimates do not show increased national influenza vaccination coverage from 2014 to 2018 in any age group.^27^ Here, we focused on adults ≥65 years old who were already covered by Medicare, and lower preventive service and premium costs under ACA may not have been substantial enough to influence the risk of influenza hospitalization.

The two analytical approaches that we have compared, mathematical modelling and statistical analysis, must each be interpreted with their strengths and limitations in mind. Mathematical models allow investigators to explicitly model transmission dynamics and offer considerable flexibility to explore a range of scenarios for parameters that cannot directly be measured, such as the distribution of prior immunity across different age groups. However, models make several simplifications in order to be tractable. In particular, mathematical models cannot explicitly model household contact structure, relying instead on contact matrices to capture inter-generational mixing. Future work could compare the results using individual-based models, which can capture household contact structure more explicitly, as well as potentially incorporating heterogeneous mixing by race/ethnicity and other groupings (data that was not available in the present study). Additionally, the uncertainty analysis of our mathematical model focused on model parameters such as prior immunity, but did not examine the effect of alternative model structures (‘structural’ uncertainty), including alternative ways of modelling vaccine-induced immunity. While statistical analyses in epidemiologic studies generally entail fewer assumptions than mathematical models, they are subject to biases of their own. Bias analysis is a useful tool for investigating the influence of potential biases but is sensitive to the assumed priors. We have higher confidence in our prior definitions for influenza testing rates since they were based on local testing rate estimates; priors for time-dependent confounding analyses were based on published studies from other populations that may not resemble values in our study population. However, our estimates of influenza testing rates included some surveillance areas that were outside of the study site, so priors may be inaccurate, and we did not have data on the relative difference in testing rates between study areas.

Our findings have several implications for future influenza vaccine studies. Prior to constructing mathematical models, it may be useful to conduct a preliminary modeling analysis to identify additional data to collect that could improve models. For example, in this study, serological analysis of biobanked, pre-intervention samples could have informed assumptions about the baseline distribution of influenza immunity within various age groups. In addition, demographic assessments could be used to construct contact matrices that account for heterogeneous mixing by race/ethnicity and other groupings. Data collection to inform potential bias analyses is also highly valuable (e.g., influenza testing rates among hospitalized patients by study group).

More generally, our approach can serve as a template for future infectious disease studies aiming to explain different estimates of intervention impact using different methods. There are many such examples, including the wide range of differing estimates of the impact of non-pharmaceutical interventions for COVID-19 from transmission models and empirical epidemiologic studies.^28^ To give another example, a transmission model and a randomized trial produced different effect estimates for combined a water, sanitation, and hygiene and deworming intervention to reduce soil-transmitted helminth infections.^29,30^

No empirical epidemiologic study can definitively rule out bias, and no mathematical model can guarantee representation of true population transmission dynamics. Systematic, quantitative investigation of the influence of model assumptions and potential biases can have equal benefits for improving both modelling and epidemiological analysis by improving future data collection and study design.

## Data Availability

All data produced are available online at: https://github.com/jadebc/SLIV-modeling-bias

## Acknowledgements

The authors would like to thank Pam Daily from the California Emerging Infections Program for her helpful input on the prior distributions for the bias-correction analysis focused on influenza testing rates among hospitalized patients. This study was supported by the Flu Lab (https://theflulab.org/) through a grant (Award number: 20142281, PI: AR) awarded to the University of California, Berkeley and by the National Institute of Allergy and Infectious Diseases of the National Institutes of Health under Award Number K01AI141616 (PI: Jade Benjamin-Chung). Jade Benjamin-Chung is a Chan Zuckerberg Biohub Investigator. Nimalan Arinaminpathy acknowledges funding from the MRC Centre for Global Infectious Disease Analysis (reference MR/R015600/1), jointly funded by the UK Medical Research Council (MRC) and the UK Foreign, Commonwealth & Development Office (FCDO), under the MRC/FCDO Concordat agreement and is also part of the EDCTP2 programme supported by the European Union; and acknowledges funding by Community Jameel. The content is solely the responsibility of the authors and does not necessarily represent the official views of the National Institutes of Health. Decisions regarding study design, data collection, statistical analysis, manuscript preparation, and publication were made independently from funders.

## Supplement 1. Supporting technical information for the mathematical model

### Specification of the mathematical model

Governing equations for the deterministic, compartmental model are as follows (see Supplementary table S1 for all parameter definitions):

Susceptible to infection, unvaccinated in the current season (*S*):

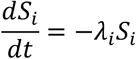

Susceptible to infection, but vaccinated in the current season (*V*_*i*_):

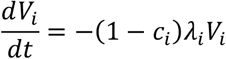

Infected and infectious, asymptomatic (*A*):

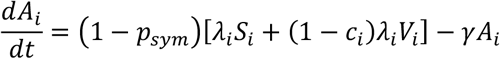

Infected and infectious, symptomatic (*I*):

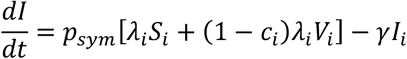

Recovered (*R*):

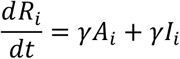

Force of infection (λ):

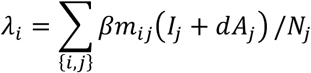

where *N*_*j*_ = *S*_*j*_ + *V*_*j*_ + *A*_*j*_ + *I*_*j*_, i.e. the total number of individuals in age group *j*, and *m*_*ij*_ is a contact matrix representing the frequency of contacts between age groups *i* and *j*, drawn from.^1^

For initial conditions, we first constructed a disease-free equilibrium as follows:

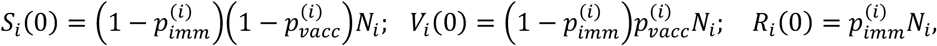

and all other state variables being zero. We then initiated an epidemic by introducing *x* infected individuals in this population, a parameter to be calibrated. The precise value of *x* does not model projections for the impact of vaccination, because it mainly serves to drive the timing of the epidemic.

### Model calibration

Free parameters in the model include: the risk of infection per contact; the proportions of each age group that are initially susceptible to infection (as determined by pre-epidemic, pre-vaccination immunity); and the rate of recovery. We also incorporated uncertainty in model parameters such as the proportion of infections that are symptomatic, and terms in the age-specific mixing matrix. Denoting these parameter values together as a vector *θ*, we defined the logarithm of the posterior density *π*(*θ*) as:

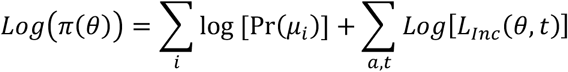

Where *μ*_*i*_ is the case-to-hospitalization multiplier; Pr(*μ*_*i*_) is the prior distribution for *μ*_*i*_ (constructed by fitting log-normal distributions to the multipliers estimated by CDC.^2^); and *L*_*Inc*_(*θ, t*) is the likelihood for observed, virologically confirmed hospitalizations in week *t* given parameters *θ*. We constructed the likelihood as follows: if the model-projected symptomatic incidence in week *t* and age group *i* is *M*_*i*_(*t*), we assumed that the reported cases in that week, *R*_*i*_(*t*), would follow a binomial distribution with number of trials *M*_*i*_(*t*) and probability of success 1/*μ*_*i*_. Approximating this with a normal distribution, we have:

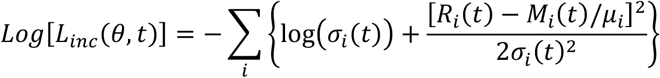

where 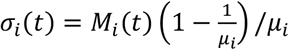, and all terms on the right-hand side of the equation above are implicitly functions of *θ*.

To sample from the posterior density, we used adaptive Bayesian Markov Chain Monte Carlo (MCMC).^3^ After drawing 10,000 samples, we discarded the ‘burn-in’ and then selected every 50^th^ sample, in order to derive 250 samples from the posterior density. We repeated this process with three different initial parameter values, sampled at random, in order to ensure convergent MCMC chains. Evaluating model projections on each of the resulting samples from the posterior density, we estimated parametric uncertainty by calculating the 2.5^th^, 50^th^ and 97.5^th^ percentiles of these model projections, denoting the interval between the upper and lower percentiles as the 95% Bayesian credible interval.

### Constrained model

To identify model parameters that would best explain differences in the results of the mathematical model and the matched cohort study, we constrained the calibration by additionally including the requirement that model projections should capture the observed reduction in hospitalization in the over-65-year-olds. In particular, we defined an alternative posterior density as:

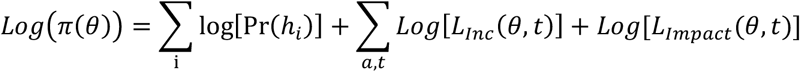

Where *L*_*Impact*_(*θ, t*) is a likelihood term for the reduction in hospitalizations amongst those over 65 years old, as a result of the intervention. We modelled *L*_*Impact*_ (*θ, t*) using a beta distribution with shape and scale parameters 37 and 99, respectively, to capture a point estimate of 22%, with 95% uncertainty intervals [15, 27].

We repeated the calibration process described above to obtain a ‘constrained’ posterior sample *θ*_*c*_. We then evaluated the ratio of unconstrained vs constrained samples, to identify any systematic differences between the two.

## Supplement 2. Prior definitions for the bias analysis for unmeasured time-dependent confounding

For P(*C*|*X*=1) and P(*C*|X=*0*), we initially defined realistic prior distributions assuming that coverage was 80% prior to the ACA (normal distribution with mean=0.8, SD = 0.01) and 90% afterwards (normal distribution with mean=0.9, SD = 0.01) based on estimates of changes in coverage in California during this time (Table S1).^4,5^

For the risk difference between hospitalization and the confounder, we defined a prior with a uniform distribution bounded by [−0.0007, 0.0007] in the pre-intervention period and bounded by [−0.0053, 0.0053] in the post-intervention period. We chose these bounds because the outcome is rare (influenza hospitalization incidence among adults 65+ years was 7 per 10,000 prior to the intervention from 2011-2014 and 53 per 10,000 during the intervention in 2017-18), so setting these bounds allowed for a percent reductions in incidence ranging from −100% to 100% at each time point. We dropped any bias-corrected case counts that were less than 0. ^6^

We also explored alternative prior distributions with less realistic assumptions to investigate what priors would be required to replicate the mathematical model’s findings. First, we investigated extreme distributions of P(*C*|*X*=1) and P(*C*|*X*=0) in which insurance coverage was approximately 0% in the intervention site (beta distribution with α = 2, β = 100) and 90% (normal distribution with mean = 0.9, SD=0.01) in the comparison site prior to the intervention and 90% in the intervention site and 0% in the comparison site during to the intervention. Second, we investigated a much stronger relationships between hospitalization and health insurance coverage, using a uniform distribution bounded by [−0.01, 0.01] both prior to and during the intervention period.

## Supplement 3. Additional tables and figures

**Table S1.**
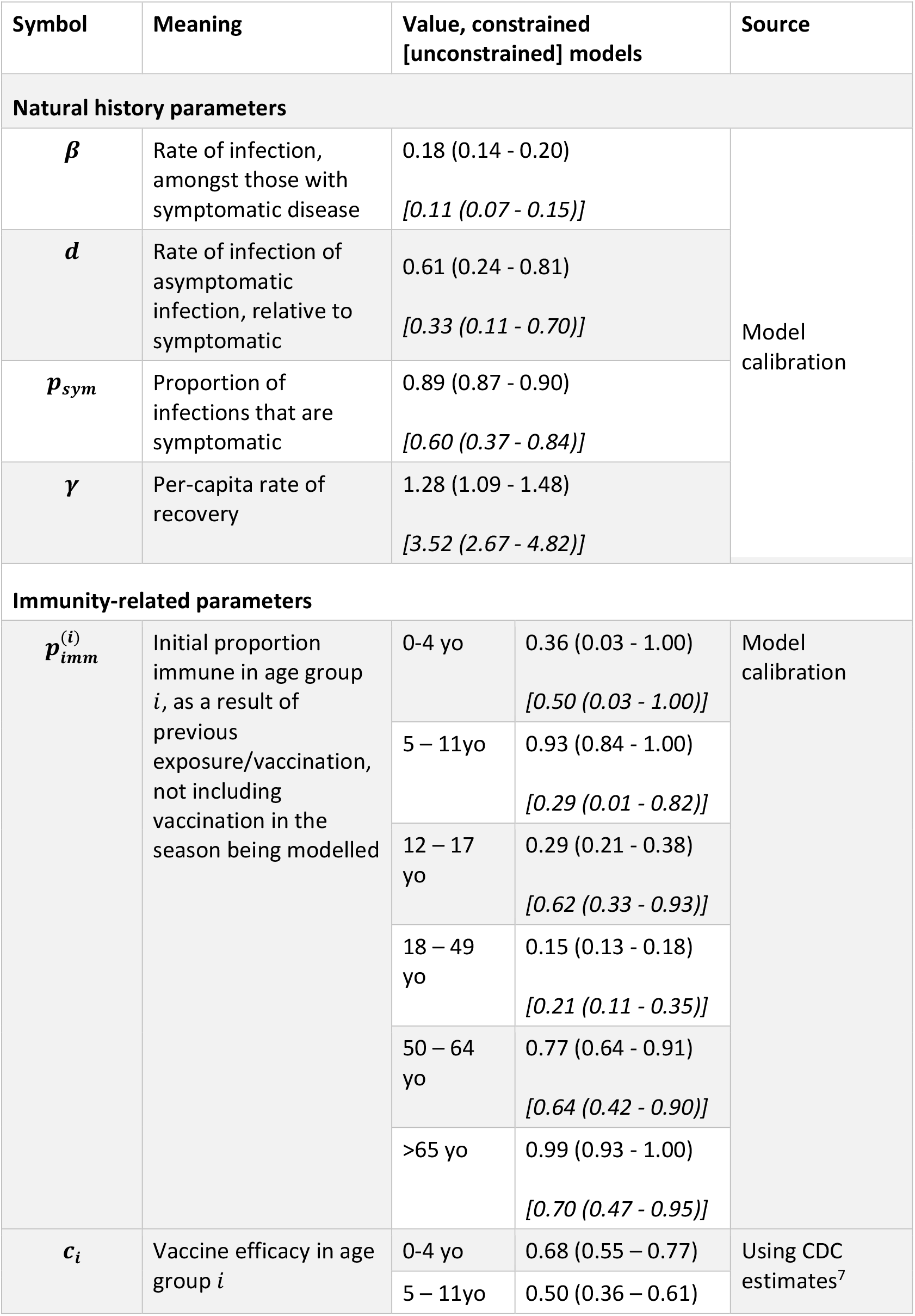
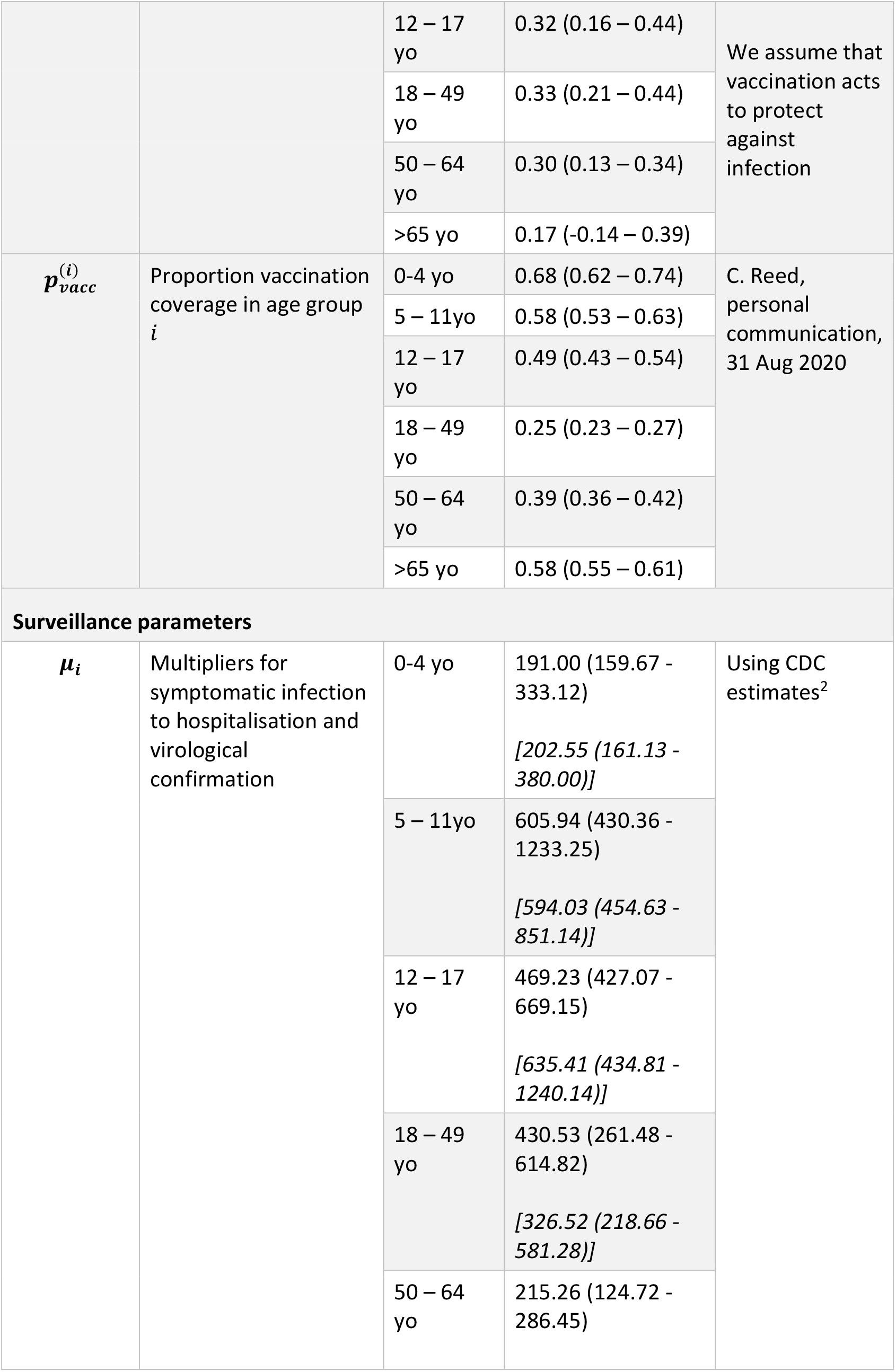
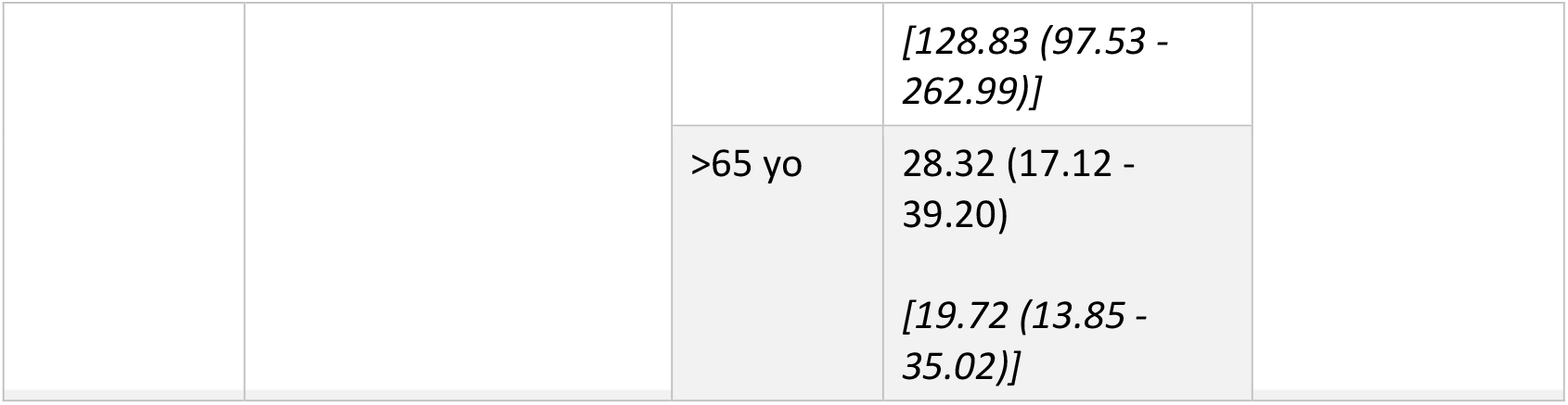
Estimates of model parameters. Numbers in round parentheses show 95% Bayesian credible interval estimates. In each cell, two sets of estimates are listed: the first is for the ‘unconstrained’ model, while the second (in square brackets) is for the ‘constrained’ model, which is constrained to capture the observed impact of augmented vaccination coverage in schoolchildren.

**Table S2.**
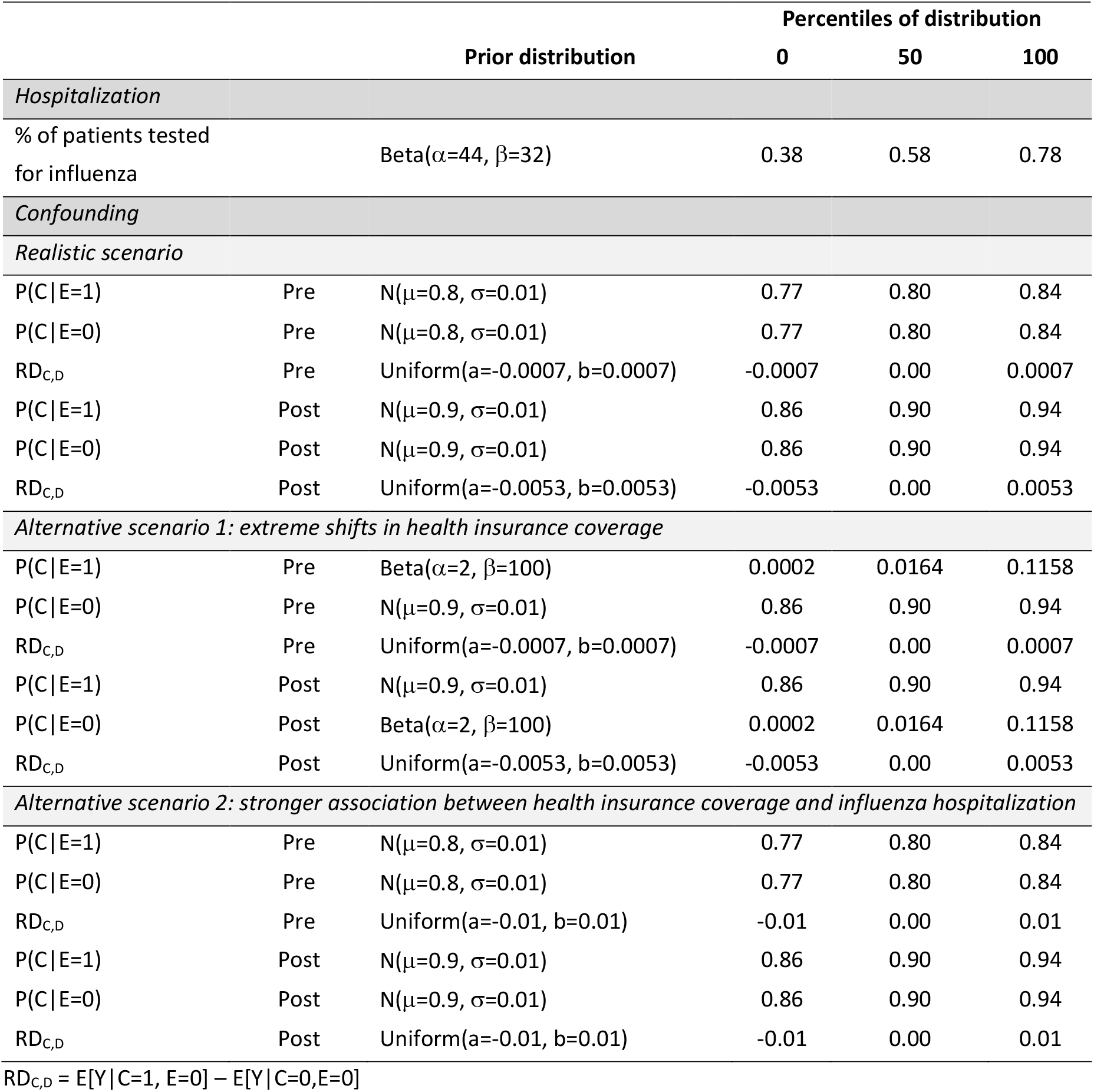
Prior distributions for probabilistic bias analysis.

**Figure S1.**
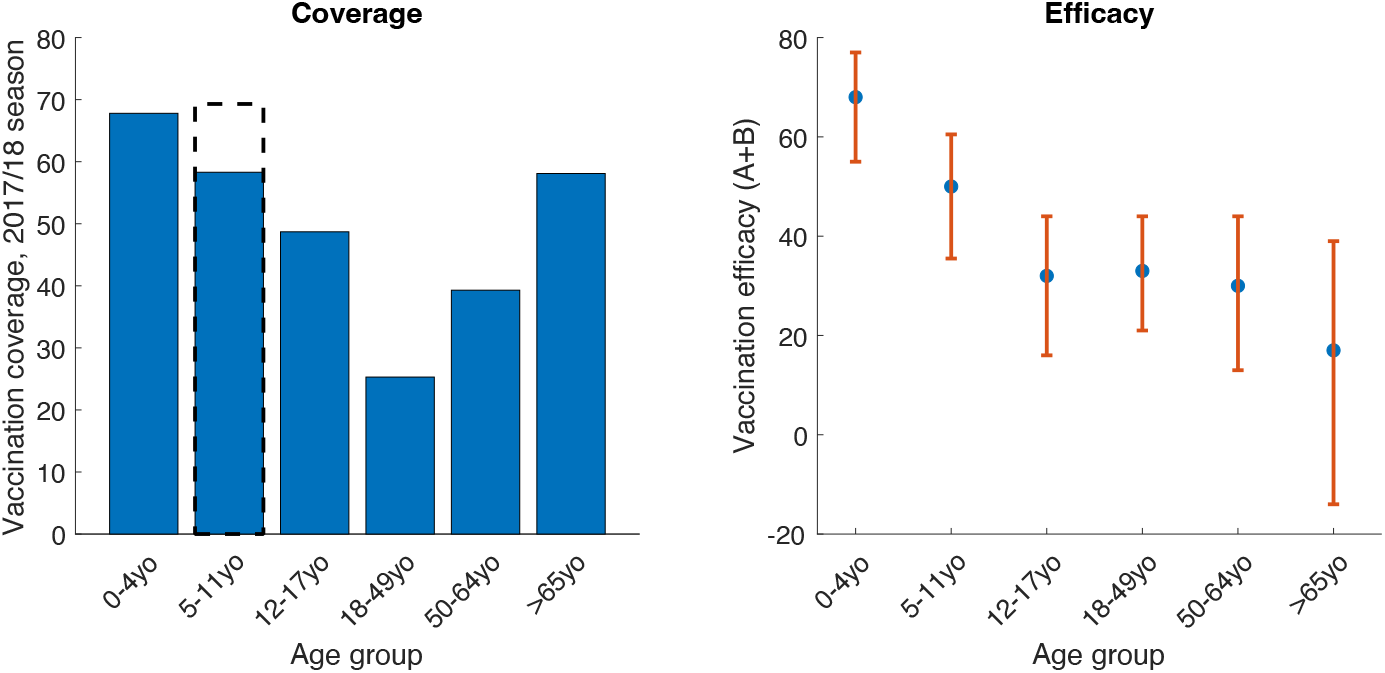
Data for vaccination coverage and efficacy used in the modelling. Left panel shows the estimates of age-specific vaccination coverage, with blue bars showing the baseline coverage (i.e. in the absence of intervention, as in the control district). These data show state-level estimates for California. The dashed black area shows the increase in vaccination coverage in the intervention district: the modelling analysis aims to capture the indirect effects of this enhancement of vaccination coverage in school children aged 5 – 11 years old. Right panel shows estimates of age-specific vaccine efficacy against influenza A in the 2017/18 season, with red lines spanning 95% uncertainty intervals.

**Figure S2.**
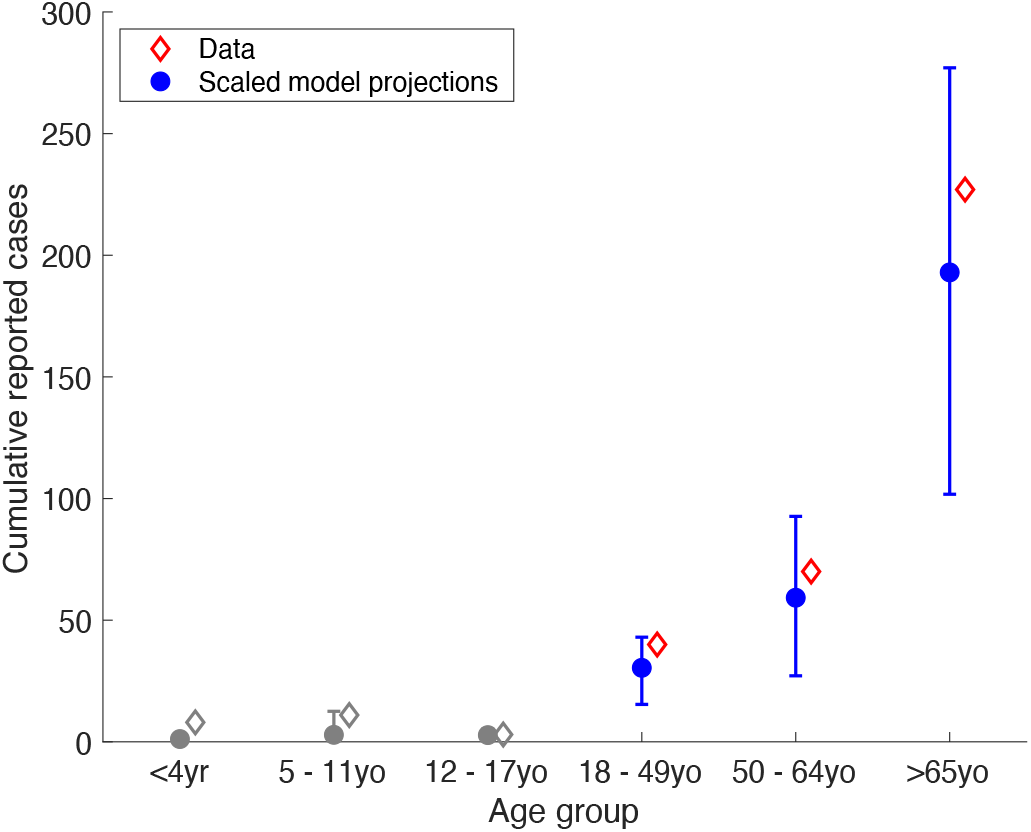
Comparison of model results and data for cumulative cases reported, for the unconstrained model. While Figure 2 in the main text shows model-data comparisons for the weekly timeseries of virologically confirmed hospitalizations, here we show comparisons for the *cumulative* reported numbers. Points for age groups lower than 18 years old are shown in grey, as these did not have sufficient data to be included as calibration targets.

**Figure S3.**
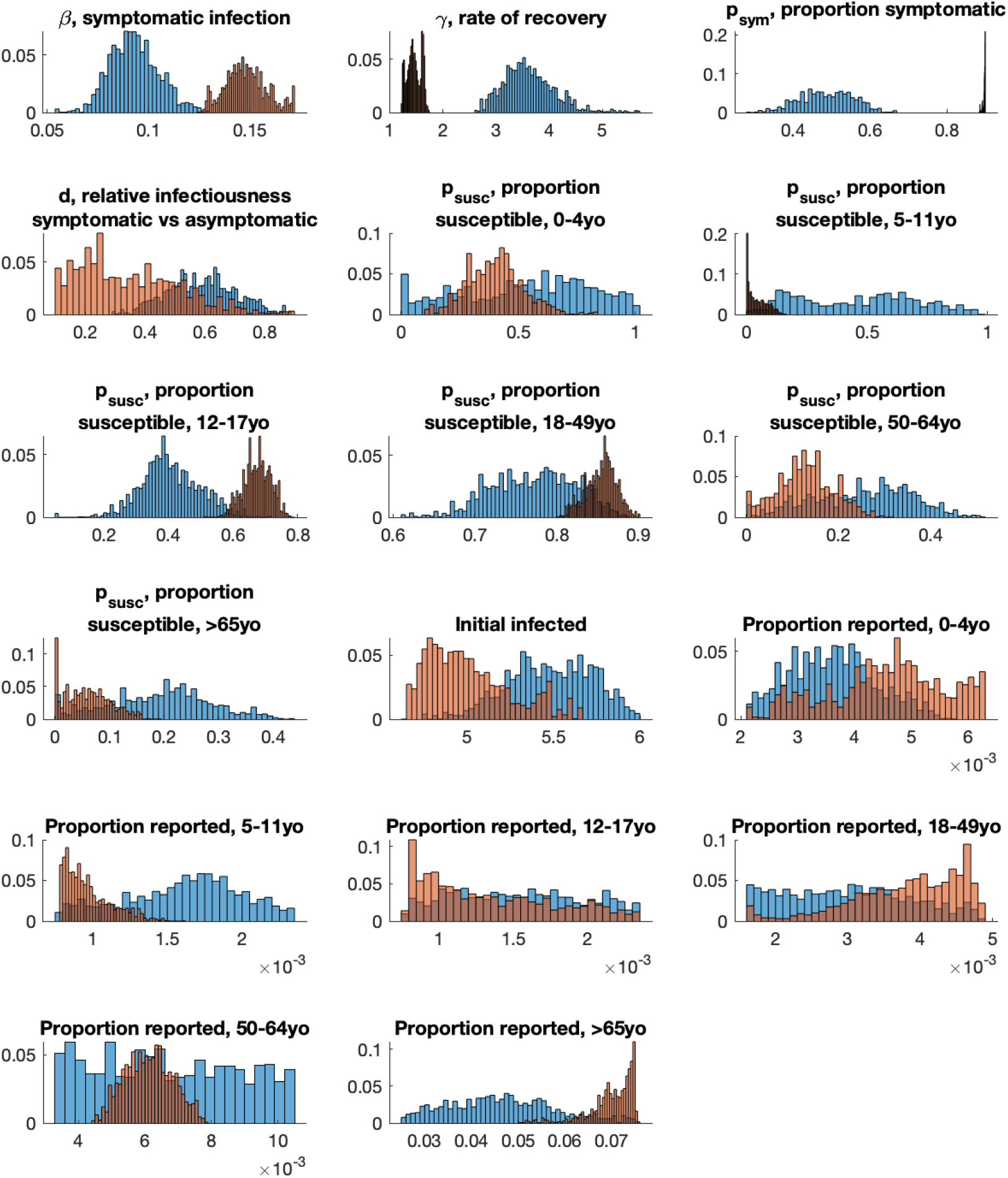
Marginal posterior densities for free parameters in the mathematical model. Histograms in blue show posterior densities for the unconstrained model, which projected a 1.6% decrease in hospitalizations amongst those over 65 years old. Histograms in orange show densities for the constrained model, which captures a 22% decrease in hospitalizations amongst those over 65 years old.

**Figure S4.**
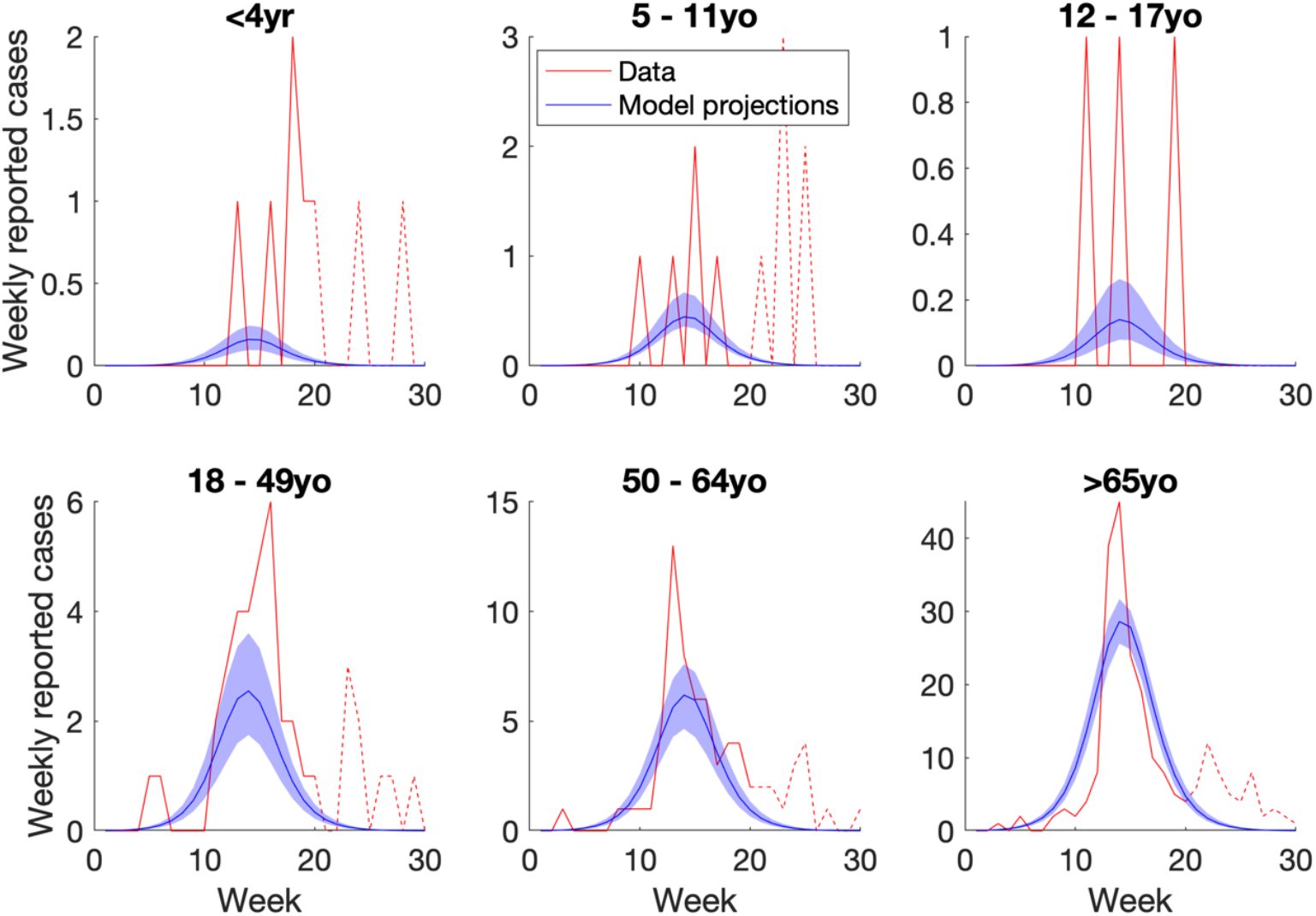
Results of calibrating the <u>constrained</u> model to epidemiological data in the control district (West Contra Costa). As for Figure 2 in the main text, curves in red show age-specific data on virologically confirmed hospitalizations from FluSURV-NET, scaled by multipliers associating this data with symptomatic incidence. Solid red lines show parts of the epidemic where influenza A dominated (to which the data was calibrated) while dashed red lines show parts driven by influenza B (not addressed in the calibration). Blue curves show best model fits, with shaded regions showing 95% Bayesian uncertainty intervals. While this Figure shows model fits to the weekly data, Figure S5 below also shows model fits to cumulative reported cases.

**Figure S5.**
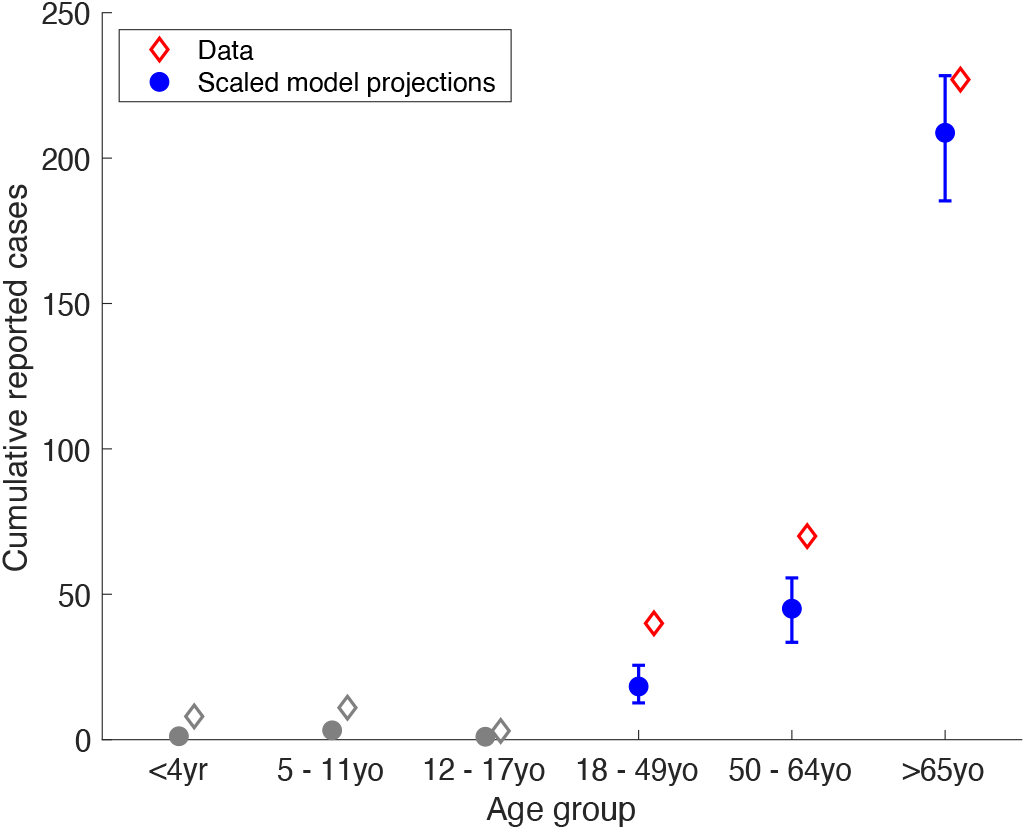
Comparison of model results and data for cumulative cases reported, for the constrained model. As for figure S2, but for the constrained model.

**Figure S6.**
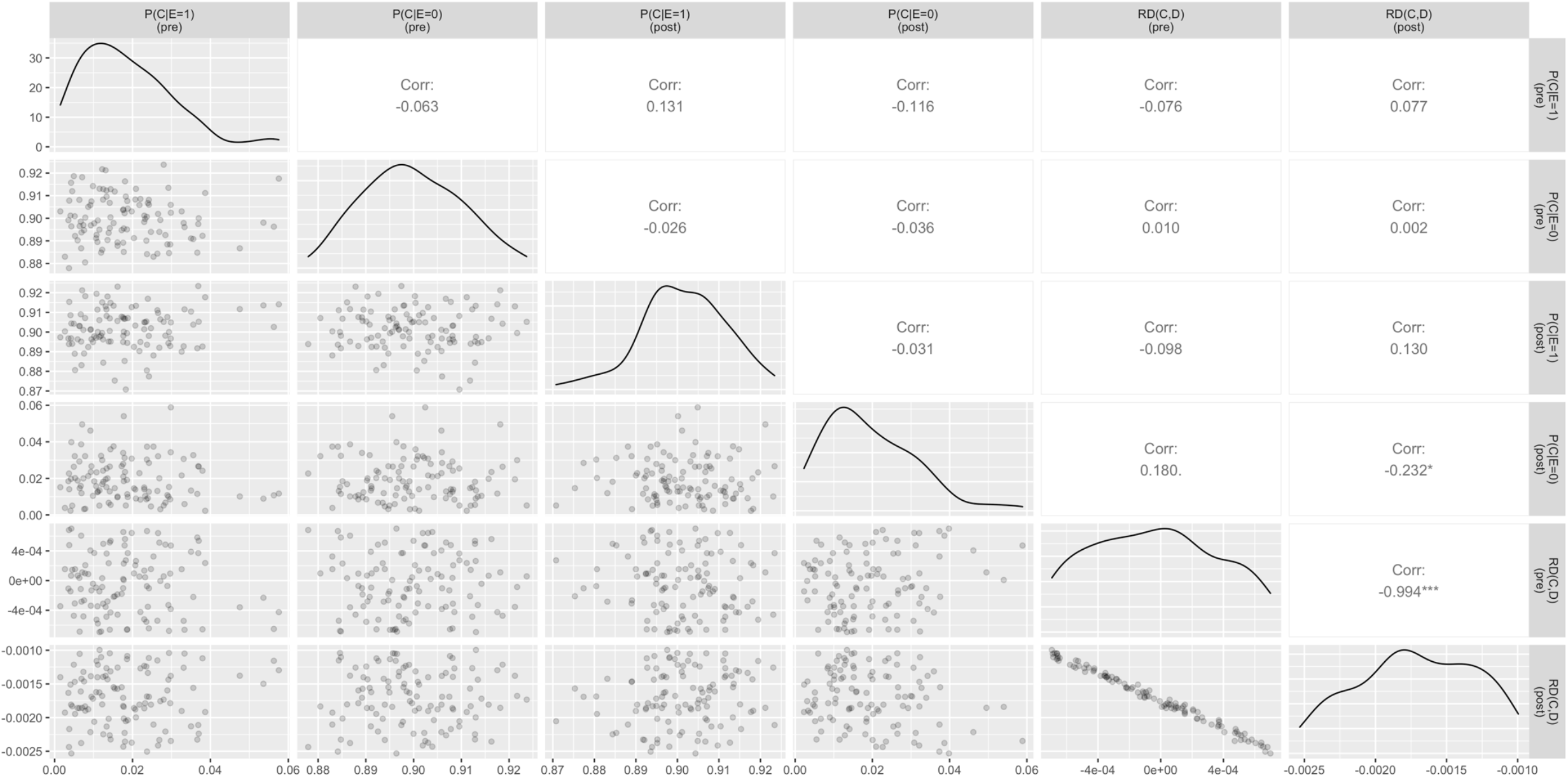
Bias-corrected estimates that matched the mathematical model. We used probabilistic bias analysis to adjust for potential time dependent confounding in the empirical matched cohort study due to the implementation of the main provisions of the Affordable Care Act, which coincided with the start of the intervention in 2014. Here, we restricted to bias-corrected estimates that matched the mathematical model. Panels on the bottom left show pairwise scatter plots of each prior. The diagonal shows univariate density distribution for each prior. Panels on the upper right show pairwise Pearson correlations.

**Figure S7.**
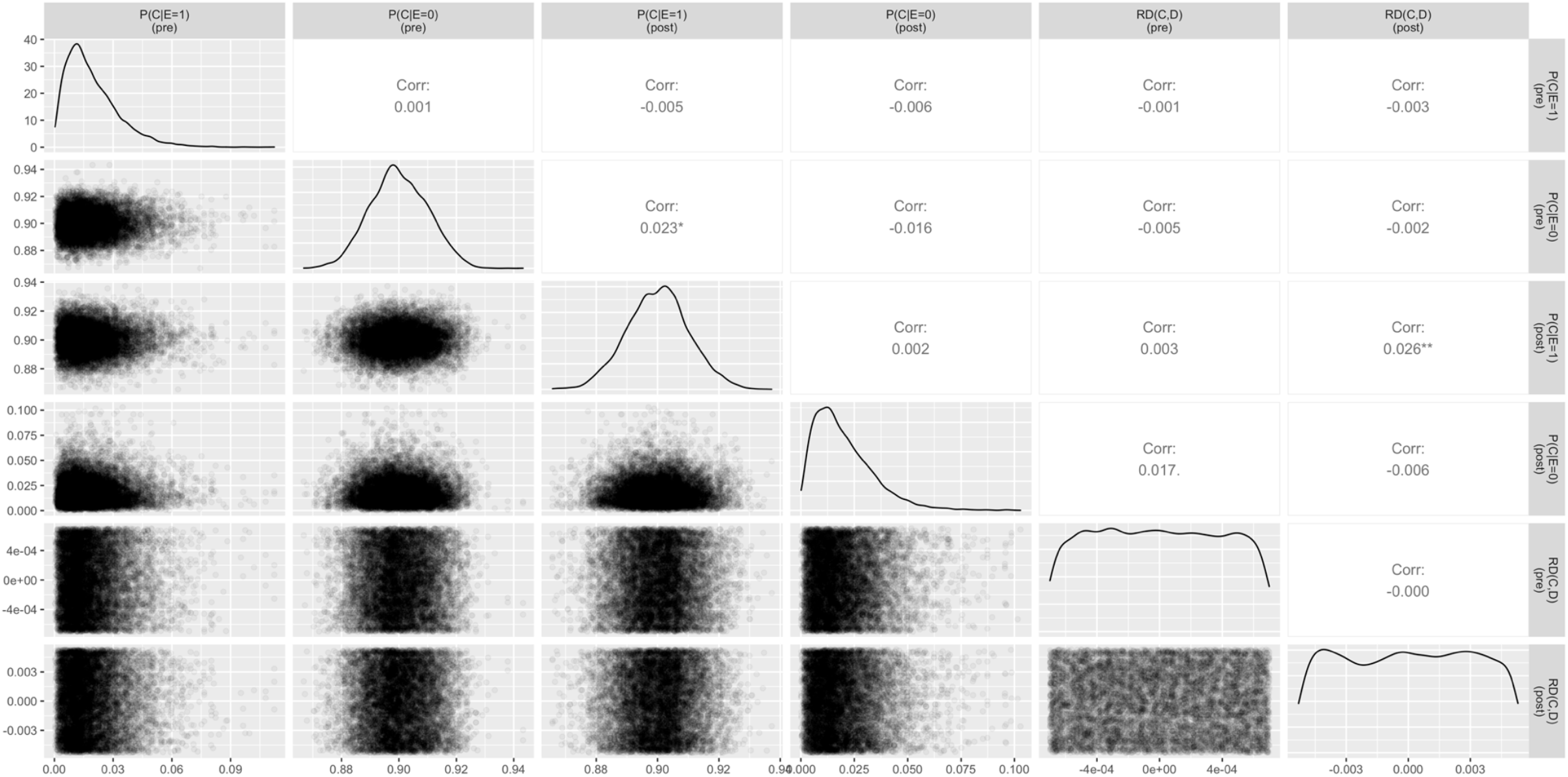
Bias-corrected estimates that did not match the mathematical model. We used probabilistic bias analysis to adjust for potential time dependent confounding in the empirical matched cohort study due to the implementation of the main provisions of the Affordable Care Act, which coincided with the start of the intervention in 2014. Here, we restricted to bias-corrected estimates that did not match the mathematical model. Panels on the bottom left show pairwise scatter plots of each prior. The diagonal shows univariate density distribution for each prior. Panels on the upper right show pairwise Pearson correlations.

## Notes

### Competing Interest Statement

The authors have declared no competing interest.

